# The COVID-related mental health load of neonatal healthcare professionals: A multicentre study in Italy

**DOI:** 10.1101/2021.06.23.21259414

**Authors:** Luigi Gagliardi, Serena Grumi, Marzia Gentile, Roberta Cacciavellani, Giulia Placidi, Angelina Vaccaro, Claudia Maggi, Beatrice Gambi, Letizia Magi, Laura Crespin, Graziano Memmini, Marcello DeFilippo, Elena Verucci, Liliana Malandra, Laura Mele, Angelo Azzarà, Livio Provenzi

## Abstract

**Background:** The COVID-19 pandemic has dramatically affected healthcare professionals’ lives. We investigated the potential mental health risk faced by healthcare professionals working in neonatal units in a multicentre cross-sectional observational study.

**Methods:** We included all healthcare personnel of 7 level-3 and 6 level-2 neonatal units in Tuscany, Italy. We measured the level of physical exposure to COVID-19 risk, self-reported COVID-related stress, and mental health load outcomes (anxiety, depression, burnout, psychosomatic, and post-traumatic symptoms) via validated, self-administered, on-line questionnaires.

**Results:** We analysed 314 complete answers. Scores above the clinical cutoff were reported by 91% of participants for anxious symptoms, 29% for post-traumatic symptoms, 13% for burnout, and 3% for depressive symptoms. Moreover, 50% of the participants reported at least one psychosomatic symptom. COVID-related stress (but not actual physical exposure) was significantly associated with all the measured mental health load outcomes, with a Risk Ratio of 3.33 (95% Confidence interval: 1.89, 5.85) for clinically relevant anxiety, 2.39 (1.69, 3.38) for post-traumatic symptoms, 1.79 (1.16, 2.75) for emotional exhaustion, and 2.51 (0.98, 6.44) for depression.

**Conclusions:** Despite a low clinical impact of COVID-19 in neonatology, neonatal professionals are a specific population at risk for psychological consequences during the pandemic.

**Keynotes:**

- We studied the mental health load (anxiety, post-traumatic, psychosomatic symptoms, burnout, depression) of healthcare professionals working in 13 neonatal units in Tuscany during the COVID-19 pandemic.
- We found very high levels of anxiety and psychosomatic symptoms, and moderate-high post-traumatic and burnout symptoms.
- Mental health load was higher in neonatal intensive (vs non-intensive) settings and in nurses (vs physicians). Mental health load outcomes were associated with COVID-related stress (rather than actual physical exposure to the virus).

## INTRODUCTION

Work-related stress is a real issue in the management of healthcare professionals. Those who are involved in complex, highly technological and/or emotionally demanding contexts are known to be at higher risk for the development of mental health problems and burnout. Physicians, nurses and other healthcare professionals who work in neonatal and pediatric contexts are required to make timely and effective decisions which – in neonatal intensive care units (NICUs) – may often concern the safety and survival of very young at-risk patients.^1,2^ Moreover, in neonatal and pediatric contexts, the healthcare providers have the double mission to provide adequate quality of care to the infants and children as well as to their parents, which may further load on their cognitive and emotional resources and may lead – in the long run – to mental health issues and psychological problems.^3,4^ Previous studies have reported high levels of mental health problems in physicians, nurses and other healthcare specialists working in pediatric and neonatal settings. In the UK, a range between 37% and 61% of physicians and nurses reported high rates of burnout^5,6^ and similar rates have been reported in Europe (36%)^7^ as well as in the United States (50%) and south America (41%).^8^

The symptoms reported by neonatal and pediatric healthcare professionals include a variety of mental health issues that may exacerbate into burnout and exhaustion. These include anxiety, depression, emotional exhaustion, psychosomatic symptoms, and post-traumatic stress.^9–15^ Compared to their colleagues working in less critical environments (e.g., neonatal wards, NWs), physicians, nurses and other healthcare professionals who work in NICUs have been found to report more difficulties in managing the emotional and psychological issues related to their job. For example, in recent investigations up to 50% of NICU professionals have reported burnout symptoms due to workload and continuous experience of a physical stressful environment.^16^ A recent study suggests that neonatal healthcare professionals may exhibit a profile of neuroendocrine dysregulation, with a flattened circadian rhythm of the hypothalamic-pituitary-adrenal axis.^17^

During the 2020, the unprecedented COVID-19 pandemic has dramatically affected the mental health of citizens^18–21^ and healthcare professionals.^22–26^ Research to date has highlighted the relevance of COVID-related stress for the worsening of psychological symptoms in professionals who care for COVID-19 patients at the frontline of the healthcare emergency. Indeed, a systematic review and meta-analysis documented a high prevalence of stress, anxiety and depression beyond the clinical risk threshold.^23^ Nonetheless, the potential traumatic consequences of the pandemic and of the related alteration of quality of life due to necessary mitigation strategies have been observed even in citizens and professionals who do not deal directly with severely ill COVID-19 patients. For example, a recent review including 62 studies from 17 countries reported a prevalence of 33% and 28% for anxiety and depression respectively, among general population and healthcare workers.^24^

The effects of the COVID-19 pandemic for healthcare professionals working in NWs and NICUs have been poorly investigated to date. Though the clinical impact of COVID-19 in perinatology has been lower than in other medical specialties, healthcare professionals have seen their working routines profoundly overhauled, often in arbitrary ways.^27^ As recently suggested,^9^ acute and chronic mental health problems in neonatal and pediatric healthcare professionals may lead to relevant impairment of the ability of the professionals to provide adequate quality of care for both the little patients and their families. The consequences may include diminished work effectiveness, decreased quality of care, poor communication with families and less efficient decision making.^7^ From this point of view, the COVID-19 pandemic may be putting an additional toll on the psychological well-being of NWS and NICU professionals, resulting in a relevant mental health load with potential detrimental consequences. A recent report about neonatal healthcare workers’ well-being showed a significant worsening, with more than the 60% of the sample exhibiting emotional exhaustion and only a third of it reporting sufficient institutional strategies to meet these emotional challenges.^28^

It is important for both professionals and healthcare systems to adequately measure and report on this mental health risk to promote appropriate preventive and therapeutic actions. As such, between May and December 2020 we launched the “Staff and Parental Adjustment to COVID-19 Epidemics – Neonatal Experience in Tuscany” (SPACE-NET) survey, with the aim of documenting the potential mental health risk faced by neonatal and pediatric healthcare professionals in Italy. In the present study we report on the mental health load experienced by physicians, nurses and other healthcare professionals who work in neonatal and pediatric NWs and NICUs.

## METHODS

### Participants and procedures

The SPACE-NET project is a multicentre cross-sectional observational study that included 7 level-3 neonatal units (that is, units that provide care including intensive care to newborns <32 weeks gestation or < 1500 g birth weight) in Tuscany, and all 6 level-2 neonatal units (that provide care to infants ≥32 weeks or > 1500 g, and no intensive care) of AUSL Toscana Nord Ovest. All the healthcare professionals working in NWs and NICUs were contacted by email. Those who participated in the survey provided an informed consent and anonymously filled in a series of questionnaires aimed at assessing their emotional stress response to the COVID-19 healthcare emergency as well as a series of potential mental health outcomes including emotional exhaustion (burnout), depression, anxiety, psychosomatic and post-traumatic symptoms. The study has been approved by the Ethics Committee of the participant parties.

### Measures

#### Socio-demographics

The socio-demographic and professional characteristics collected were sex, age (years), setting (NW or NICU), job (physician, nurse, or other, including psychologists, social workers, residents, midwives, physiotherapists, rehabilitation technicians, and auxiliary healthcare assistants), and job experience (years).

#### COVID-19 exposure and COVID-related stress response

The participants’ direct (own infection or risk of infection) or indirect (infection or risk of infection of significant others) physical exposure to SARS-CoV2 was assessed with an ad-hoc 5-item questionnaire previously used by our group.^29^ The items of this questionnaire were rated dichotomously. A global index was obtained by summing all the responses and re-coding the sum into a *COVID-19 exposure* variable coded as “no” if sum was equal to 0 or “yes” if sum was above 0. The COVID-related stress response was assessed using an ad-hoc 6-item questionnaire previously used by our group.^30^ The items were rated 1 (low stress) to 5 (high stress). A mean global index was computed and used in this study as *COVID-related stress index*. The items included in *COVID-19 exposure* and *COVID-related stress index* are reported in Supplementary File S1.

#### Mental health outcomes

The symptoms related to the following domains of mental health status were investigated: depression, anxiety, psychosomatic symptoms, emotional exhaustion, and post-traumatic symptoms. Depressive symptoms were measured using the Beck Depression Inventory – II (BDI-II)^31^, a 21-item scale widely used to assess subclinical and clinical depressive symptomatology. Each BDI-II item is rated on a 4-point Likert scale and a global sum score (*depressive symptoms*) is obtained with severe depression indexed by scores higher than 28. Anxious symptoms were rated using the state anxiety subscale of the State-Trait Anxiety Inventory – Y form (STAI-Y).^32^ The state anxiety subscale features 20 4-point Likert items that are summed up in global score (*anxious symptoms*); scores higher than 40 indicate a risk for clinically relevant anxiety. A list of psychosomatic symptoms obtained from the Psychosomatic Symptom Checklist^33^ was rated by each participant for severity on a 6-point Likert scale (1 = low, 6 = high). A mean score was obtained to index *psychosomatic symptoms*. A subscale of the Maslach Burnout Inventory (MBI)^34^ measuring *emotional exhaustion* was also included. This subscale includes nine 7-point Likert scale items (1 = low, 7 = high). A global *emotional exhaustion* score is obtained by summing the items ratings and it indexes clinical symptoms if higher than 30. The Impact of Event Scale (IES)^35^ was used to assess the *post-traumatic symptoms*. The IES is a 22-item questionnaire. Each item is rated on a 5-point Likert scale and a global sum score is obtained; scores higher than 33 are meant to suggest the presence of clinical risk.

### Statistical analysis

Univariate analyses were carried out using analysis of variance (ANOVAs) or independent-sample *t*-tests. The linear association between *COVID-related stress index* and mental health outcomes was assessed by means of Pearson’s bivariate correlations. For variables for which a validated clinical severity cutoff is available (depression, anxiety, burnout, post-traumatic symptoms), a dichotomous variable [above cutoff/below cutoff] was computed, and binomial regressions were used to estimate the probability of scoring above cutoff, including the following predictors in the model: setting, job, job experience, and *COVID-related stress index*.

As all mental health domains were correlated, in order to reduce the number of statistical comparisons and obtain an overall index, we used a principal component analysis (PCA) to calculate a global *mental health load index* (*MHLI*) that would explain the largest portion of variance in mental health outcomes (i.e., *depressive symptoms, anxious symptoms, psychosomatic symptoms, post-traumatic symptoms*, and *emotional exhaustion*). For this analysis we set the minimum Eigenvalue to 1 and we adopted a non-rotated solution. We used the principal component with the highest loading and that explained the highest portion of variance as the primary outcome variable for the study. The MHLI has mean = 0 and standard deviation = 1.

Specific mental health outcomes were further tested as additional endpoints. The statistical analyses were conducted using R^36^ and IBM SPSS Statistics for Windows, ver. 26.0.^37^

## RESULTS

A total of 314 healthcare professionals participated in the study, out of 941 invited (32.9%). The majority were females (n = 281, 89.5%) and working as physicians (n = 100, 31.8%) or nurses (n = 145, 46.2%), 192 were working in NWs (61.1%) whereas the remaining 122 (38.5%) were working in NICUs. About half of the sample had a direct or indirect exposure to COVD-19, including a 10.5% who experienced the death of a friend or significant person. Healthcare professionals in NICUs reported higher *COVID-related stress index* compared to NW counterparts, *(p* = .014). Descriptive statistics are reported in Table 1.

**Table 1.**
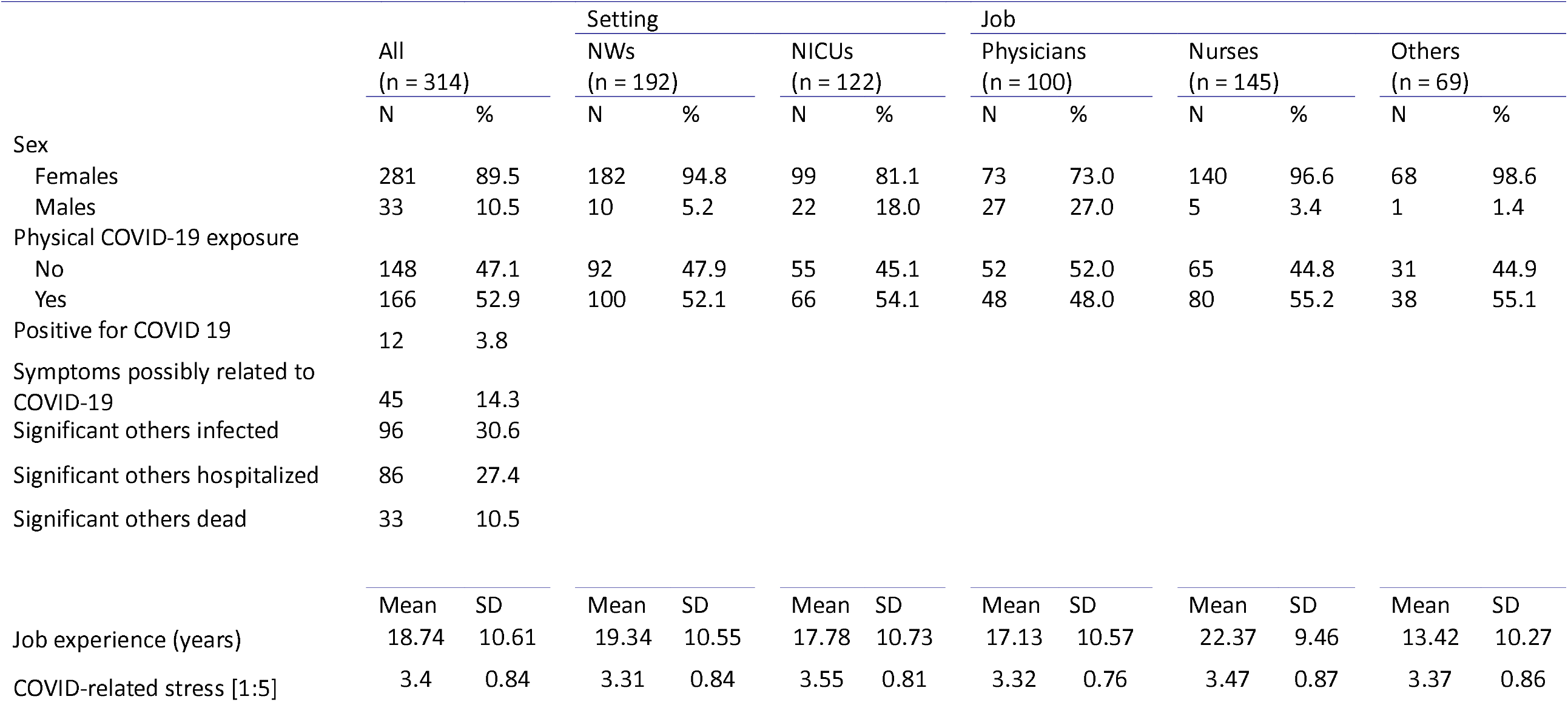
Descriptive statistics of the studied sample.

The mental health status in the various domains investigated are shown in Table 2. Nurses reported higher *anxious symptoms* when compared to physicians, but not to other professionals, *p* = .041. Scores above the clinical cutoff were reported by 91% of participants for *anxious symptoms*, 29% of participants for *post-traumatic symptoms*, 13% of participants for *emotional exhaustion*, and 3% of participants for *depressive symptoms*. Moreover, 50% of the participants reported at least one psychosomatic symptom.

**Table 2:**
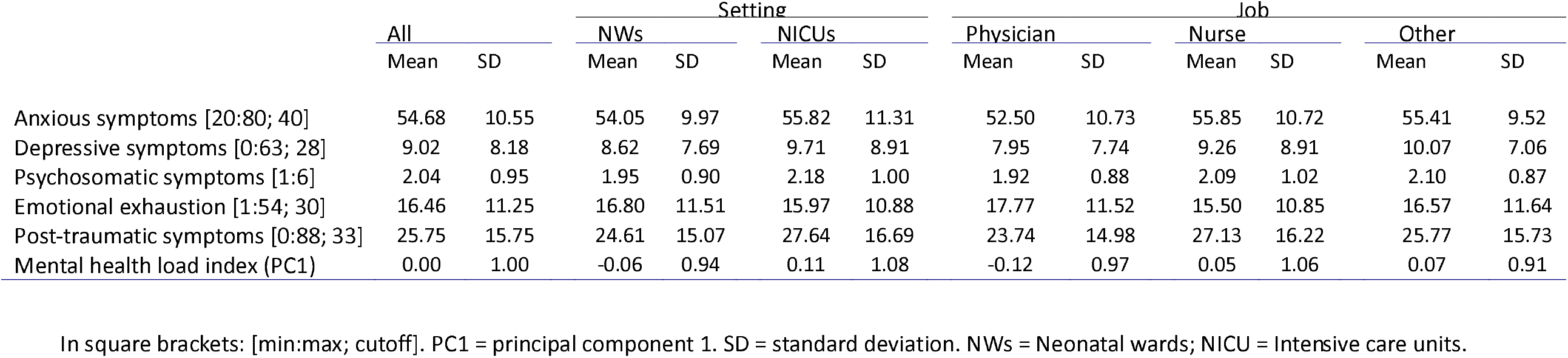
Mental health outcomes in the studied sample

The principal component analysis yielded a one-component solution, the *mental health load index*, explaining the 65.3% of total variance and with loadings ranging from 0.67 to 0.90. No statistically significant differences in *MHLI* emerged for job and setting. *COVID-19 exposure* was not correlated with *MHLI* nor with any specific mental health load outcomes. *COVID-related stress index* significantly correlated with all mental health load outcomes as well as with the *MHLI* (Figure 1). Figure 2 reports the association between *COVID-related stress index* and *MHLI* by setting and job.

**Figure 1.**
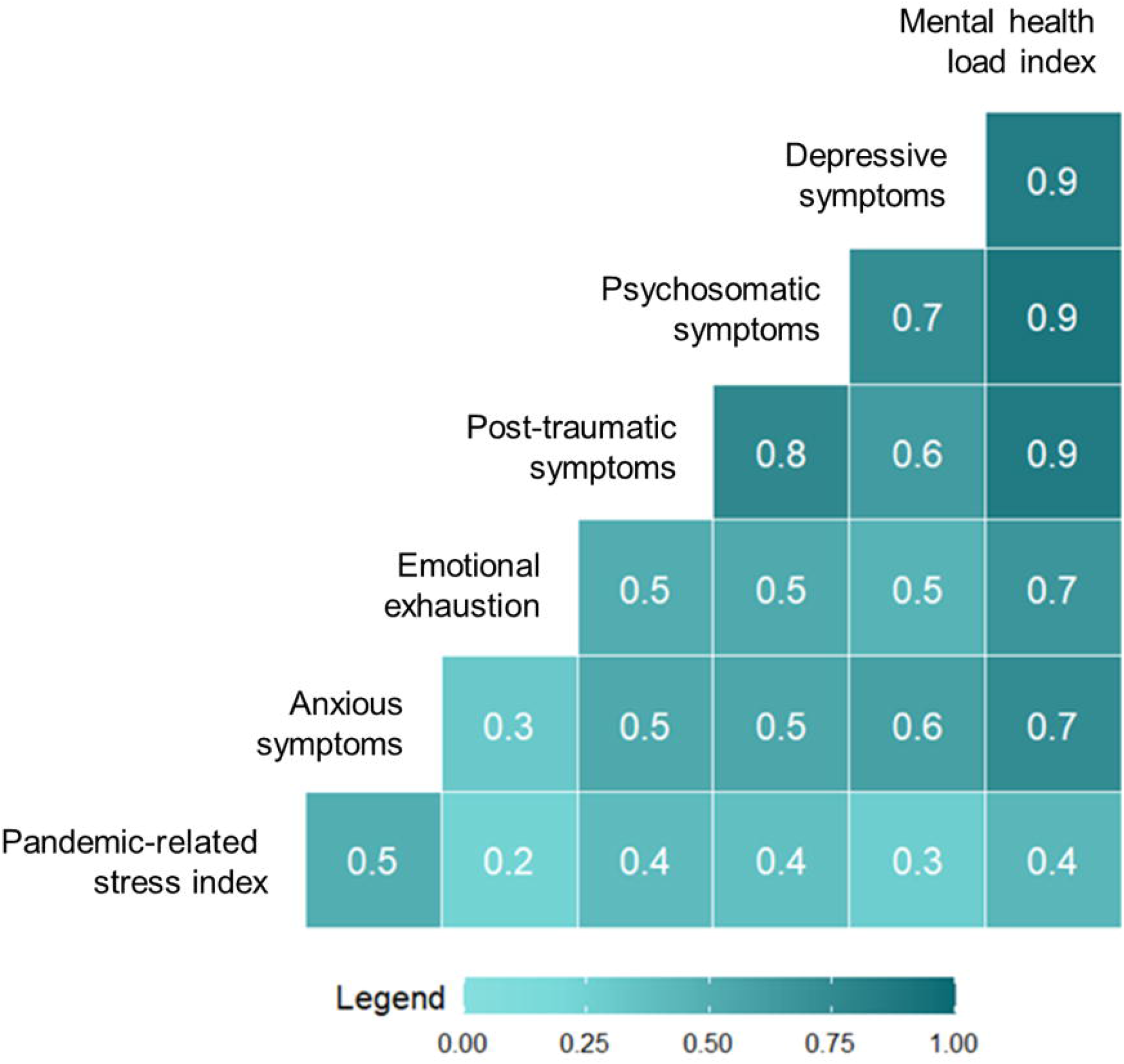
Bivariate correlations between COVID-related stress and mental health outcomes.

**Figure 2.**
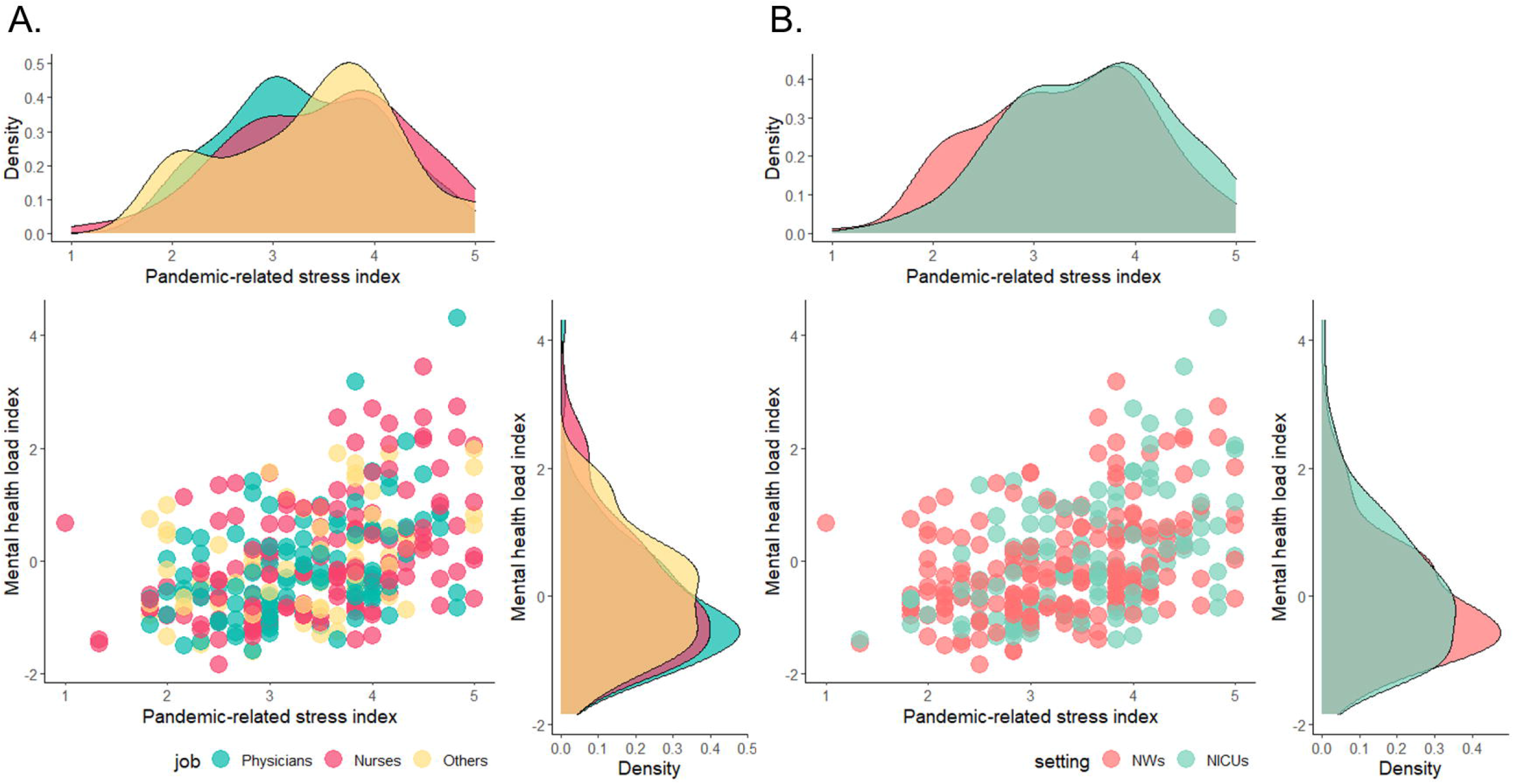
Association between COVID-related stress and mental health load index split by job (A) and setting (B).

Binomial regressions (Table 3) showed that an increase of one point in *COVID-related stress index* was significantly associated with an increased risk of clinically relevant (i.e., above the accepted cutoff level) *anxious symptoms* (RR = 3.33; 95% CI 1.89, 5.85), *post-traumatic symptoms* (RR = 2.39; 95% CI 1.69, 3.38), and clinically relevant *emotional exhaustion (RR = 1*.*79, 95%CI* 1.16, 2.75). The results for depressive symptoms bordered statistical significance (*RR* = 2.51, *95%CI* 0.98, 6.44). No significant effects of setting, job, and job experience emerged for any of the outcomes.

**Table 3.** Association of COVID-related stress index with clinically significant outcomes. Binomial regression models.

| A. Anxious symptoms |  |  |  |
| --- | --- | --- | --- |
|  | *RR* | 95% CI | *p* |
| COVID-related stress index | 3.33 | [1.89;5.85] | < .001 |
| Setting (NWs) |  |  | Reference |
| Setting (NICUs) | 0.53 | [.21;1.34] | 0.18 |
| Job (physician) |  |  | Reference |
| Job (nurse) | 2.32 | [.92;5.86] | 0.08 |
| Job (other) | 1.27 | [.36;4.45] | 0.71 |
| Job experience | 0.98 | [.94;1.02] | 0.29 |
| B. Post-traumatic symptoms |  |  |  |
|  | *RR* | 95% CI | *p* |
| COVID-related stress index | 2.39 | [1.69;3.38] | < .001 |
| Setting (NWs) |  |  | Reference |
| Setting (NICUs) | 1.55 | [.86;2.80] | 0.15 |
| Job (physician) |  |  | Reference |
| Job (nurse) | 1.16 | [.61;2.19] | 0.65 |
| Job (other) | 1.14 | [.55;2.34] | 0.73 |
| Job experience | 1.00 | [.97;1.03] | 0.92 |
| C. Emotional exhaustion |  |  |  |
|  | *RR* | 95% CI | *p* |
| COVID-related stress index | 1.79 | [1.16;2.75] | 0.01 |
| Setting (NWs) |  |  | Reference |
| Setting (NICUs) | 0.60 | [.27;1.33] | 0.21 |
| Job (physician) |  |  | Reference |
| Job (nurse) | 0.52 | [.23;1.17] | 0.12 |
| Job (other) | 1.03 | [.43;2.45] | 0.96 |
| Job experience | 1.01 | [.98;1.05] | 0.41 |
| D. Depressive symptoms |  |  |  |
|  | *RR* | 95% CI | *p* |
| COVID-related stress index | 2.51 | [.98;6.44] | 0.06 |
| Setting (NWs) |  |  | Reference |
| Setting (NICUs) | .85 | [.19;3.81] | 0.21 |
| Job (physician) |  |  | Reference |
| Job (nurse) | 5.43 | [.59;9.65] | 0.13 |
| Job experience | .99 | [.91;1.07] | 0.73 |

RR: Risk Ratio; NWs: neonatal wards; NICU: neonatal intensive care units; SE: standard error. Job (other) not included in the D model, as no healthcare professionals in this subgroup reported significant depressive symptoms above the clinical cutoff.

## DISCUSSION

The present study aimed at investigating the mental health load experienced by physicians, nurses and other healthcare professionals who work in NWs and NICUs during the COVID-19 pandemic. In our sample, more than 90% of participants reported anxious symptoms above the clinical cutoff, half of participants experienced at least one psychosomatic symptom and about one third of the sample reported a post-traumatic symptomatology above the clinical risk. These results suggest that also professionals in the perinatal field experienced the increased emotional burden documented for physicians working at the forefront of the pandemic during the COVID-19 emergency.^18^ Moreover, the severity of COVID-related stress largely impacted on their psychophysical health. Though very few professionals had had COVID-19, more than half of the sample experienced a direct exposure to the disease, including 10.5% who experienced the death of a significant other, and 27.4% a hospital admission. Thus, it is not unexpected that even if these professionals were not directly involved in the care of patients positive for COVID-19, these results were comparable to those reported for an Italian sample of frontline healthcare professionals.^18^ Anxious symptoms especially emerged to be the more reactive outcome when facing critical situations, representing a sort of red flag of professionals’ mental health, perhaps also caused by the profound overhaul of established professional daily tasks.

Differently from the first published report on burnout experienced by NWs and NICUs workers,^28^ in our Italian context a limited percentage of NWs and NICUs professionals exhibited a level of emotional exhaustion compatible with a full-blown burnout. The effects of COVID-19-related stress on the depressive symptomatology were apparently limited, suggesting that depression may be a less reactive outcome during emergency crisis.^38^ Nonetheless, it should be highlighted that we used the more restrictive BDI-II cutoff, which indicates the presence of severe depressive signs. It is also necessary to highlight the cross-sectional nature of the study that did not allow to disentangle the potential impact of COVID-related stress from that of the usual workload. Therefore, it is not possible to exclude that the reported depressive symptoms may be a carry-over effect of difficulties related to the pre-pandemic period.

Physicians and nurses showed the same levels of physical COVID-19 exposure, independently of the type of unit (NW or NICU), while significant differences in COVID-related stress and anxiety emerged for job and setting. In particular, professionals of NICUs exhibited a higher COVID-19-related stress that may be linked to the greater impact of containment measures on their professional practices. As for job, nurses exhibited higher levels of anxiety.^39^ This difference emerged also in previous studies performed during the pre-pandemic period, showing a higher anxiety for nurses – especially if working in intensive units – linked to their work-related activities.^39^ Therefore, this higher vulnerability may have been exacerbated during the lockdown period.

Our study has limitations. Firstly, the response rate of our survey (32.9%), though similar to that of another recent study,^28^ does not allow us to claim representativeness of our sample. Secondly, the cross-sectional study design does not allow to assess the causal directions of the relationship between the COVID-related stress and the professionals’ wellbeing. Moreover, the unavailability of pre-emergency data did not allow to disentangle the potential impact of COVID-related stress from that of the usual workload. Although data collection occurred by self-report questionnaires, we used well-validated tools, except for the ad-hoc measure used to assess COVID-19 exposures and response. Finally, participants were enrolled from hospitals located in only one Italian region that was not a primary hotspot of the virus spreading during the first lockdown, but that was hit during subsequent waves of the pandemic,^40^ as confirmed by the 10% of respondents who had experienced the death of at least one significant other and 30% of them who had indirect experience of a relative or close friend who needed intensive care hospitalization.

## Conclusions

The COVID-19 pandemic is likely to exert a relevant stress toll on the mental health of neonatal and pediatric healthcare professionals. Even if they are not directly involved in the care of patients positive for COVID-19, they should be considered as a specific population at risk for psychological consequences of the pandemic. As such, appropriate actions are needed from clinical institutions and policymakers to mobilize dedicated resources to take care of their psychological health.

## Data Availability

Due to stipulations between participating hospitals, these data are not publicly available.
Motivated requests can be addressed to the corresponding author.

## Contributors’ statement

LG and LP conceived the study. SG, RC and GP made substantial contributions to the design of the work. MG, AV, CM, BG, LMag, LC, GM, MDF, EV, LMal, LMe, AA organized and coordinated the acquisition of data for the work. SG and LP performed the analysis and interpretation of the data. LG, LP and SG drafted the manuscript. All the authors revised it critically for important intellectual content, and

## REFERENCES

1. Bucher HU, Klein SD, Hendriks MJ, et al. Decision-making at the limit of viability: Differing perceptions and opinions between neonatal physicians and nurses. BMC Pediatr. 2018;18(1). doi:10.1186/s12887-018-1040-z

2. Kadivar M, Mosayebi Z, Asghari F, Zarrini P. Ethical challenges in the neonatal intensive care units: Perceptions of physicians and nurses; an iranian experience. J Med Ethics Hist Med. 2015;8. https://pubmed.ncbi.nlm.nih.gov/26839675/. Accessed April 18, 2021.

3. Latour JM, Hazelzet JA, Duivenvoorden HJ, Van Goudoever JB. Perceptions of parents, nurses, and physicians on neonatal intensive care practices. J Pediatr. 2010;157(2). doi:10.1016/j.jpeds.2010.02.009

4. Mirlashari J, Brown H, Fomani FK, de Salaberry J, Zadeh TK, Khoshkhou F. The Challenges of Implementing Family-Centered Care in NICU from the Perspectives of Physicians and Nurses. J Pediatr Nurs. 2020;50:e91–e98. doi:10.1016/j.pedn.2019.06.013

5. Colville G, Dalia C, Brierley J, Abbas K, Morgan H, Perkins-Porras L. Burnout and traumatic stress in staff working in paediatric intensive care: associations with resilience and coping strategies. Intensive Care Med. 2015;41(2):364–365. doi:10.1007/s00134-014-3559-2

6. Colville G. Paediatric intensive care nurses report higher empathy but also higher burnout than other health professionals. Evid Based Nurs. 2018;21(1):25. doi:10.1136/eb-2017-102774

7. Rodríguez-Rey R, Palacios A, Alonso-Tapia J, et al. Burnout and posttraumatic stress in paediatric critical care personnel: Prediction from resilience and coping styles. Aust Crit Care. 2019;32(1):46–53. doi:10.1016/j.aucc.2018.02.003

8. Fields AI, Cuerdon TT, Brasseux CO, et al. Physician burnout in pediatric critical care medicine. Crit Care Med. 1995;23(8):1425–1429. doi:10.1097/00003246-199508000-00018

9. Crowe S, Sullivant S, Miller-Smith L, Lantos JD. Grief and burnout in the PICU. Pediatrics. 2017;139(5). doi:10.1542/peds.2016-4041

10. Dryden-Palmer K, Garros D, Meyer EC, Farrell C, Parshuram CS. Care for Dying Children and Their Families in the PICU: Promoting Clinician Education, Support, and Resilience. Pediatr Crit Care Med. 2018;19(8S Suppl 2):S79–S85. doi:10.1097/PCC.0000000000001594

11. Favrod C, du Chêne LJ, Soelch CM, et al. Mental health symptoms and work-related stressors in hospital midwives and NICU nurses: A mixed methods study. Front Psychiatry. 2018;9(AUG). doi:10.3389/fpsyt.2018.00364

12. Hynan MT, Steinberg Z, Baker L, et al. Recommendations for mental health professionals in the NICU. J Perinatol. 2015;35(S1):S14–S18. doi:10.1038/jp.2015.144

13. Lima-Setta F, Pacheco D, Stochero L, et al. P0315 / #1683: Prevalence of anxiety and depression in health professionals working in pediatric intensive care units in Rio De Janeiro/Brazil. Pediatr Crit Care Med. 2021;22(Supplement 1 3S):171–171. doi:10.1097/01.pcc.0000739600.12091.0d

14. Lin TC, Lin HS, Cheng SF, Wu LM, Ou-Yang MC. Work stress, occupational burnout and depression levels: A clinical study of paediatric intensive care unit nurses in Taiwan. J Clin Nurs. 2016;25(7-8):1120–1130. doi:10.1111/jocn.13119

15. Meadors P, Lamson A. Compassion Fatigue and Secondary Traumatization: Provider Self Care on Intensive Care Units for Children. J Pediatr Heal Care. 2008;22(1):24–34. doi:10.1016/j.pedhc.2007.01.006

16. Bresesti I, Folgori L, De Bartolo P. Interventions to reduce occupational stress and burn out within neonatal intensive care units: a systematic review. Occup Environ Med. 2020;77(8):515–519. doi:10.1136/oemed-2019-106256

17. Fumagalli M, Provenzi L, Sorrentino G, et al. Self-Report and Biological Indexes of Work-Related Stress in Neonatal Healthcare Professionals: A Repeated-Measures Observational Study. Adv Neonatal Care 2021. https://europepmc.org/article/med/33538493. Accessed April 18, 2021.

18. Barello S, Palamenghi L, Graffigna G. Burnout and somatic symptoms among frontline healthcare professionals at the peak of the Italian COVID-19 pandemic. Psychiatry Res. 2020;290(May):113129. doi:10.1016/j.psychres.2020.113129

19. Xiong J, Lipsitz O, Nasri F, et al. Impact of COVID-19 pandemic on mental health in the general population: A systematic review. J Affect Disord. 2020;277:55–64. doi:10.1016/j.jad.2020.08.001

20. Wang Y, Kala MP, Jafar TH. Factors associated with psychological distress during the coronavirus disease 2019 (COVID-19) pandemic on the predominantly general population: A systematic review and metaanalysis. PLoS One. 2020;15(12 December). doi:10.1371/journal.pone.0244630

21. Salari N, Hosseinian-Far A, Jalali R, et al. Prevalence of stress, anxiety, depression among the general population during the COVID-19 pandemic: A systematic review and meta-analysis. Global Health. 2020;16(1). doi:10.1186/s12992-020-00589-w

22. da Silva FCT, Neto MLR. Psychological effects caused by the COVID-19 pandemic in health professionals: A systematic review with meta-analysis. Prog Neuro-Psychopharmacology Biol Psychiatry. 2021;104. doi:10.1016/j.pnpbp.2020.110062

23. Salari N, Khazaie H, Hosseinian-Far A, et al. The prevalence of stress, anxiety and depression within front-line healthcare workers caring for COVID-19 patients: a systematic review and meta-regression. Hum Resour Health. 2020;18(1). doi:10.1186/s12960-020-00544-1

24. Luo M, Guo L, Yu M, Wang H. The psychological and mental impact of coronavirus disease 2019 (COVID-19) on medical staff and general public – A systematic review and meta-analysis. Psychiatry Res. 2020;291. doi:10.1016/j.psychres.2020.113190

25. Kisely S, Warren N, McMahon L, Dalais C, Henry I, Siskind D. Occurrence, prevention, and management of the psychological effects of emerging virus outbreaks on healthcare workers: rapid review and meta-analysis. BMJ. 2020;369:m1642. doi:10.1136/bmj.m1642

26. Trumello C, Bramanti SM, Ballarotto G, et al. Psychological adjustment of healthcare workers in italy during the COVID-19 pandemic: Differences in stress, anxiety, depression, burnout, secondary trauma, and compassion satisfaction between frontline and non-frontline professionals. Int J Environ Res Public Health. 2020;17(22):1–13. doi:10.3390/ijerph17228358

27. Yeo KT, Oei JL, De Luca D, et al. Review of guidelines and recommendations from 17 countries highlights the challenges that clinicians face caring for neonates born to mothers with COVID-19. Acta Paediatr. 2020 Nov;109(11):2192-2207. doi: 10.1111/apa.15495

28. Haidari E, Main EK, Cui X, et al. Maternal and neonatal health care worker well-being and patient safety climate amid the COVID-19 pandemic. J Perinatol. March 2021. doi:10.1038/s41372-021-01014-9

29. Grumi S, Provenzi L, Gardani A, et al. Rehabilitation services lockdown during the COVID-19 emergency: the mental health response of caregivers of children with neurodevelopmental disabilities. Disabil Rehabil. 0(0):1–6. doi:10.1080/09638288.2020.1842520

30. Provenzi L, Grumi S, Giorda R, et al. Measuring the Outcomes of Maternal COVID-19-related Prenatal Exposure (MOM-COPE): Study protocol for a multicentric longitudinal project. BMJ Open. 2020;10(12). doi:10.1136/bmjopen-2020-044585

31. Beck AT, Steer RA, Carbin MG. Psychometric properties of the Beck Depression Inventory: Twenty-five years of evaluation. Clin Psychol Rev. 1988;8(1):77–100. doi:10.1016/0272-7358(88)90050-5

32. Spielberger, C. D. State-Trait Anxiety Inventory: Bibliography. 1989, Palo Alto, CA: Consulting Psychologists Press.

33. Attanasio V, Andrasik F, Blanchard EB, Arena JG. Psychometric properties of the SUNYA revision of the psychosomatic symptom checklist. J Behav Med. 1984;7(2):247–257. doi:10.1007/BF00845390

34. Maslach C, Leiter MP, Schaufeli W. Measuring Burnout. In: The Oxford Handbook of Organizational Well Being. Oxford University Press; 2009. doi:10.1093/oxfordhb/9780199211913.003.0005

35. Horowitz M, Wilner N, Alvarez W. Impact of Event Scale: a measure of subjective stress. Psychosom Med. 1979;41(3):209–218.

36. R Core Team. R: A language and environment for statistical computing. R Foundation for Statistical Computing, 2014, Vienna, Austria.

37. IBM Corp. IBM SPSS Statistics for Windows, version 27

38. Kendler KS, Karkowski LM, Prescott CA. Stressful life events and major depression: Risk period, long-term contextual threat, and diagnostic specificity. J Nerv Ment Dis. 1998;186(11):661–669. doi:10.1097/00005053-199811000-00001

39. Nooryan K, Gasparyan K, Sharif F, Zoladl M. Controlling anxiety in physicians and nurses working in intensive care units using emotional intelligence items as an anxiety management tool in Iran. Int J Gen Med. 2012;5:5–10. doi:10.2147/IJGM.S25850

40. Tuscany Region. [COVID-19 data in Tuscany and Italy]. Retrieved from: https://www.ars.toscana.it/banche-dati/dati-sintesi-sintcovid-aggiornamenti-e-novita-sul-numero-dei-casi-deceduti-tamponi-per-provincia-e-per-asl-della-regione-toscana-e-confronto-con-italia-con-quanti-sono-i-decessi-per-comune?provenienza=home_ricerca&dettaglio=ric_geo_covid&par_top_geografia=090

